# The Use of Saliva as a Diagnostic Specimen for SARS CoV-2 Molecular Diagnostic Testing for Pediatric Patients

**DOI:** 10.1101/2020.11.11.20223800

**Authors:** Meghan Delaney, Joelle Simpson, Bobbe Thomas, Christal Ralph, Michael Evangalista, Mahdi Moshgriz, Joyce Granados, Mark McGuire, Roberta DeBiasi, Joseph Campos

## Abstract

**Background:** Children are an important population to test for COVID-19 infection, particularly because they may shed the virus without displaying symptoms. Testing children for COVID-19 via sensitive molecular methods is important, although collecting nasopharyngeal (NP) specimens can be challenging. A less invasive mode of specimen collection that yields test results comparable to those from NP specimens would be beneficial to simplify sample collection.

**Methods:** To demonstrate that saliva is a suitable specimen for collection from children, the clinical usability/acceptability and the analytic performance of saliva were compared to NP specimens suspended in viral transport medium. Four different FDA EUA-approved real-time RT-PCR assays and one EUA approved saliva collection device were investigated.

**Results:** The study population included 526 patients between the ages of 3 and 61 years, 461 (88%) were <18 years, 425 were asymptomatic (81.1%), 92 were symptomatic (17.6%). Saliva mixed with saliva stabilizing buffer was found to yield comparable sensitivity to NP specimens when tested on the AllPlex SARS-CoV-2 molecular test (Seegene Inc). The analytic sensitivity of the AllPlex assay during testing of spiked saliva mixed with SpectrumDNA saliva stabilizer was found to be 250 genomic copies/mL.

**Conclusions:** Of the four FDA EUA-approved SARS-CoV-2 PCR assays studied, we found the AllPlex assay to be best suited for testing saliva specimens collected from children 5 years of age or older. The sensitivity of viral detection was equivalent to NP specimens when saliva specimens were mixed with the saliva stabilizer.

## INTRODUCTION

The COVID-19 pandemic continues to ravage the global population. Virtually every populated region of the world, with the sole exception of Antarctica, is experiencing ongoing infections.^1^ While individuals of all age groups have been shown susceptible to infection, children are less likely than adults to experience severe infections.^2,3^ Children rarely may progress to multisystem inflammatory syndrome in children (MIS-C). MIS-C is a newly described post-infectious condition that occurs in children 2 – 4 weeks after exposure to COVID-19.^4^ Children may also be asymptomatically infected.^5^ Testing children for COVID-19 via molecular methods is important because low levels of virus may be present, although collecting nasopharyngeal (NP) specimens is challenging in children, especially for repeated testing which may be used in “back to school” initiatives. To identify childhood vectors of infection and diagnose children with symptoms of the disease, an easier mode of collecting specimens would be beneficial.

The use of saliva as a diagnostic specimen for detecting SARS-CoV-2 virus in the clinical laboratory has not been straightforward. Saliva is not a typical sample type used in diagnostic clinical testing; it may have interferences and viscosity levels that makes pipet manipulation challenging. A recently published letter to the editor described an unsuccessful attempt to obtain comparably sensitive polymerase chain reaction (PCR) results from saliva and simultaneously collected NP specimens from children.^6^ The study involved testing unprocessed saliva using a laboratory-developed real time, reverse transcription polymerase chain reaction (RT-PCR) test for the SARS-CoV-2 E gene. The authors concluded that saliva is not a useful specimen for diagnosis of COVID-19 infection in children. In contrast, Wyllie et al followed a similar study design in testing saliva from adult patients and detected higher concentrations of SARS-CoV-2 RNA in self-collected saliva than in NP specimens collected at the same time by health care workers.^7^ They found that a higher percentage of saliva specimens than NP specimens were PCR positive during the first 10 days of COVID-19 infection and concluded that the testing of saliva and NP specimens had at least similar sensitivities in the detection of SARS-CoV-2 by PCR in symptomatic individuals.

We sought to establish the utility of saliva specimens for detection of SARS-CoV-2 RNA in symptomatic and asymptomatic pediatric patients. We evaluated the testing of untreated saliva as well as saliva mixed with a commercially available saliva stabilizing solution (SpectrumDNA, Spectrum Solutions, Draper, UT). The SpectrumDNA saliva collection system has been granted Emergency Use Approval (EUA) by the U.S. Food and Drug Administration (FDA) during the declared COVID-19 pandemic national emergency. SARS-CoV-2 PCR testing of processed and unprocessed saliva was performed with four EUA-approved assays: Cepheid Xpert Xpress, DiaSorin Simplexa, GenMark ePlex, and Seegene AllPlex. We also wished to understand the success of saliva collection from children, who are in different developmental stages and may have different abilities to provide a saliva specimen.

## MATERIALS & METHODS

To determine if saliva was a suitable specimen to use to test children for the presence of the SARS-CoV-2 virus, institutional review board (IRB) approval was obtained and a two-part study was undertaken. The first part of the study was designed to assess clinical usability/acceptability of saliva as a specimen source in pediatric patients. The second part of the study consisted of an analytical validation study of saliva specimens compared to specimens that were collected from the NP. During the study, saliva and NP specimens were tested with the EUA-approved RT-PCR assays available in our high complexity CLIA-accredited laboratory. The assays were evaluated with respect to validity of results, sensitivity of viral detection using PCR cycle time (CT) comparison and limit of detection (LOD) analysis as appropriate.

### Clinical usability

We conducted a prospective study of patients who were being tested for COVID-19 infection in the Emergency Department (ED), peri-operative testing program and employees being tested through occupational health at our facility. The study population included symptomatic and asymptomatic patients. Each study participant was tested for SARS-CoV-2 by nasopharyngeal (NP) or oropharyngeal (OP) swab at our institution. After informed consent was obtained, subjects were asked to not to eat or drink for 30 minutes prior to saliva collection and then each subject provided a saliva specimen. The study used two different collection devices; a urine cup to collect unprocessed saliva (target volume of 1 – 2 mL) and a commercially available EUA-approved SpectrumDNA saliva collection kit that includes a tube with a line marking a standard collection volume (3 mL) and a proprietary stabilizing buffer solution (1.5 mL) that is mixed with the saliva immediately after collection per the manufacturer’s instructions. We sought to determine whether different saliva collection containers had characteristics that would be more favorable for sample collection and testing. Study patients were queried as to whether they felt ill or well at the time of specimen collection.

We determined the age at which children were able to reliably provide saliva specimens of sufficient quantity to be successfully analyzed in the laboratory. Because expectorating on demand requires a minimal level of behavioral maturity, we wished to determine the lowest age at which adequate saliva collection could be reasonably expected, either due to behavioral readiness or by producing enough saliva for testing within a reasonable period of time.

### Analytical validity

One of the study goals was to determine which of the EUA-approved SARS-CoV-2 PCR assays employed by our laboratory could be successfully adapted to test human saliva. Available assays included the AllPlex™ 2019-nCoV Assay (Seegene, Inc., Seoul, South Korea), the ePlex SARS-CoV-2 Test (GenMark Dx, Carlsbad, CA), the Xpert Xpress SARS CoV-2, Cepheid, Sunnyvale, CA) and the Simplexa™ COVID-19 Direct assay (DiaSorin Molecular LLC, Cypress, CA)]. The FDA has stated that they do not intend to object to the use of a test, without their notification or submission of a new or amended EUA, when the test is a modification of an EUA-authorized test such as using saliva as a specimen type. Samples used in the validation studies included those obtained from the clinical usability study described above, spiked saliva specimens using remnant specimens obtained via NP swab collections and a commercially available SARS-CoV-2 Standard quantitated at 200,000 copies/mL of all PCR gene targets (Exact Diagnostics, Fort Worth, TX).

To test saliva specimens using the Simplexa assay, the manufacturer’s instructions were followed that consists of loading samples directly into the test cartridges without a preceding extraction step. The Simplexa assay targets the ORF 1ab region and the S gene of the SARS-CoV-2 genome and uses a cycle time positive to negative cut-off of 40 PCR cycles. To test saliva specimens using the AllPlex (Seegene) assay, specimens were extracted off-line using either a manual extraction system (Zymo Quick-RNA) or an automated system (QiaSymphony, Qiagen EZ1 Advanced (Qiagen, Maryland, USA)). The AllPlex assay targets specific nucleic acid sequence of the E gene, RdRP gene (R), and the N gene of the SARS-CoV-2 genome and uses a positive to negative cut-off of 40 PCR cycles. To test saliva specimens using the Xpert Xpress (Cepheid) assay, specimens were extracted and amplified within the test cartridge. The Xpert Xpress assay targets specific nucleic acid sequences of the E gene and the N gene of the SARS-CoV-2 genome and uses a positive to negative cut-off of 45 PCR cycles. To test saliva specimens using the ePlex (GenMark) assay, samples were extracted within the test cartridge. The ePlex test system does not provide a CT value as part of the result. Because the testing of unprocessed saliva with the ePlex assay revealed complete inhibition of amplification of the internal control, we elected to discontinue further evaluation of the assay for saliva specimens.

All four assays were tested with unprocessed saliva and all but the ePlex assay were tested with saliva collected in using the SpectrumDNA saliva collection kit. When invalid results (e.g. failure of the internal control to amplify) were obtained, the saliva specimen was diluted 1:1 with viral transport medium and retested. In addition, when an invalid result was obtained with the AllPlex assay, specimens were also re-extracted using an alternative extraction method (either manual or automated as described above). Further manipulations to obtain valid results included Proteinase-K and heat pretreatment as described by others.^8^

A limit of detection (LOD) analysis of the AllPlex assay was performed starting with a SARS-CoV-2 commercial standard containing 200,000 copies/mL of all SARS-CoV-2 gene targets (Exact Diagnostics). Ten-fold serial dilutions of the standard were prepared down to 1,000 copies/mL followed by two-fold serial dilutions down to 125 copies/mL to obtain a more precise LOD. Twenty replicates of the dilutions containing 500 copies/mL and 250 copies/mL were tested to determine the concentration at which at least 19 of the replicates yielded positive results. In all tests, if one or more genes were detected, the result was considered positive.

## RESULTS

The study was undertaken during July – September 2020 at Children’s National Hospital which is an urban, pediatric, tertiary care medical center located in Washington, DC. The hospital primarily serves children living in the Washington, DC metropolitan area, including northern Virginia, and Maryland. The study population included 526 patients between the ages of 3 and 61 years. The patients in the study had an overall positive test result rate of 2.4% (12 of 510 valid results). Of the patients, 461 (88%) were <18 years and 425 were asymptomatic (81.1%), 92 were symptomatic (17.6%) and the symptoms were not recorded in nine patients. Ten samples did not have enough volume for testing and were excluded (10 of 526, 1.9%). The unacceptable samples were collected from individuals throughout the age range, with more seen in younger subjects giving unprocessed saliva (Figure 1). The collecting staff found that children five years of age and older had the developmental maturity to consistently provide satisfactory saliva specimens, although younger children generally required more time than older children and adolescents. There were 425 asymptomatic subjects (81.1%), 92 subjects with symptoms suggestive of COVID-19 infection (17.6%) and nine subjects for whom symptomology information was not recorded.

**Figure 1.**
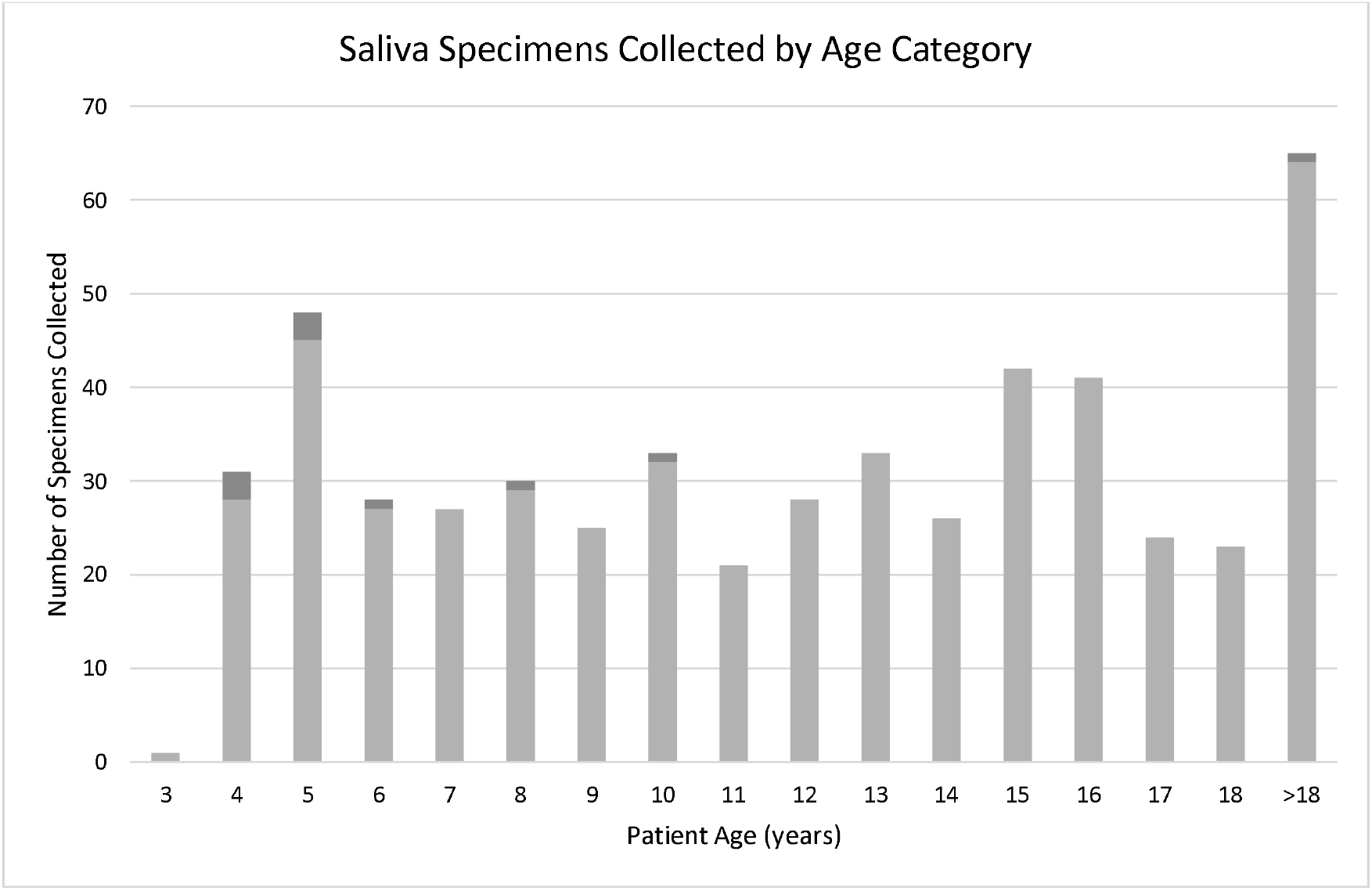
The acceptability to use saliva specimen collection in children. Light gray bars are the number of specimens successfully collected; dark gray bars are the number of specimens that were insufficient quantity for testing.

The Simplexa (Diasorin) assay had 52 initially invalid samples (14.2% of 365) due to failed internal controls. Almost all of the samples with invalid test results yielded valid results after diluting the samples with an equal volume of viral transport medium (51 of 52, 98.1%). Saliva samples spiked with positive NP specimens had higher CT values compared to the NP specimen result (data not shown). When unprocessed saliva samples were mixed with the SpectrumDNA saliva stabilizer or treated with Proteinase-K and heat, the Simplexa assay internal controls did not amplify. Accordingly, we decided to discontinue further testing with the Simplexa assay for clinical use.

The Xpert Xpress (Cepheid) assay testing was limited by the number of test cartridges we had available. Three examples of saliva spiked with positive NP specimens and diluted in the SpectrumDNA saliva stabilizer provided valid positive results. Saliva samples spiked with positive NP specimens had higher CT values compared to the NP specimen result (data not shown), although this can be partially attributed to the dilution. Because of the low inventory of the test cartridges, we did not move forward with further testing and limit of detection analysis.

The Allplex (Seegene) assay had 23 initially invalid samples (9.8% of 235 samples) due to failed internal controls. None of the samples with invalid test results yielded valid results after diluting the samples with an equal volume of viral transport medium. However, when saliva specimens were mixed with the SpectrumDNA saliva stabilizer there were zero invalid results. Spiking 30 saliva samples is SpectrumDNA saliva stabilizer with 100 uL of positive NP specimen yielded 100% correlation of positive results with comparable CT values (Figure 2a-c). There were five simultaneously collected positive NP specimens and SpectrumDNA saliva positive samples tested on the AllPlex assay (Figure 2d-f). There were no negative saliva results when the matched NP specimen was positive. The analytic sensitivity of the Allplex assay of saliva in SpectrumDNA saliva stabilizer was determined by calculating the limit of detection (LOD). The LOD was found to be 250 viral RNA copies/mL.

**Figure 2.**
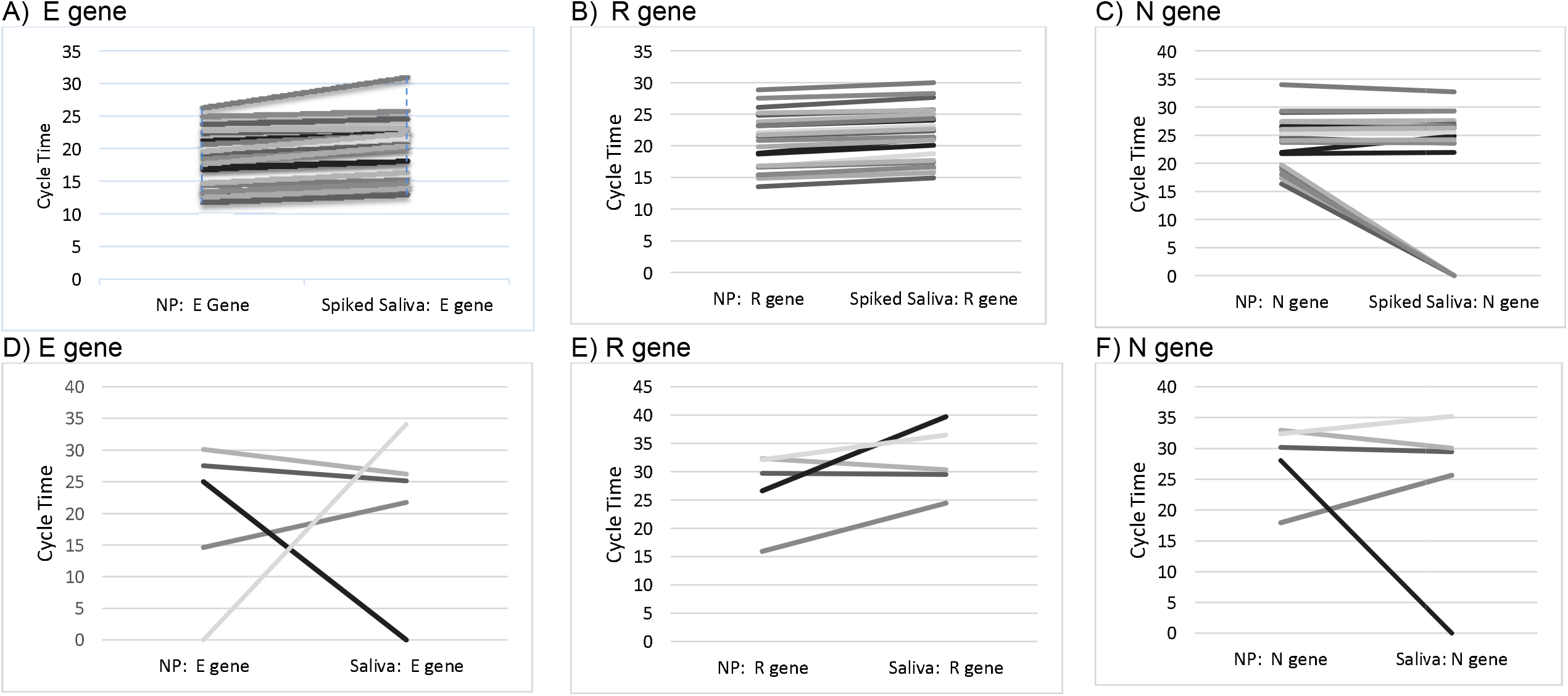
Comparison of nasopharyngeal (NP) swab and saliva specimens tested by RT-PCR for three gene targets in the Allplex (Seegene) assay. Panels A – C used a 100uL aliquot of each positive NP sample into 100uL of negative saliva collected in the SpectrumDNA collection kit, creating a 1:1 dilution. Panels D – F shows NP specimens and contemporaneous saliva specimens were collected using the specimen collection device (SpectrumDNA) in close time proximity. These results meet the recommended criterion of at least 95% positive and negative agreement for acceptable clinical performance.

We concluded that the AllPlex assay of saliva samples collected with the SpectrumDNA saliva collection kit provided the optimum combination of favorable features including excellent analytic sensitivity, ease of use and results comparable to those obtained from testing NP samples. We established the recommended lower age limit for collection of saliva samples with the SpectrumDNA saliva collection kit to be five years based on the experience of our collecting staff. This age recommendation is not based on analytical performance.

## DISCUSSION

During this investigation, we wished to determine whether saliva specimens could substitute for NP specimens when testing symptomatic and asymptomatic children old enough to produce sufficient amounts of saliva upon demand. Our experience suggests an age cut-off of five years of age, although some younger children were able to successfully provide saliva. Theoretically, PPE-protected suctioning of saliva could be employed with younger children, although we did not pursue that line of investigation during our study.

We assessed the compatibility of four EUA-approved SARS-CoV-2 PCR assays for testing unprocessed saliva. Our findings indicated that for three of the four assays we were able to detect SARS-CoV-2 virus using saliva (Cepheid Xpert Xpress, DiaSorin Simplexa, and Seegene AllPlex), but the PCR cycle thresholds (CTs) comparison, the internal control and/or number of invalid samples and/or availability of test cartridges was not consistent enough to be suitable for ongoing clinical use. Using saliva mixed with the SpectrumDNA saliva stabilizer demonstrated successful amplification of the internal controls for the AllPlex assay and had comparable sensitivity (CT values) to the NP specimens. Because of the availability of PCR reagents and specimen testing throughput we performed a LOD analysis to determine the analytic sensitivity of the AllPlex assay. The results demonstrated an LOD of 250 genomic copies/mL, a result consistent with, or better than the LOD available from the manufacturer (1250 – 4160 viral copies/mL).^9^

Our study has some limitations. Although over 500 patients gave a saliva specimens as part of this study, the number with a positive PCR test result was low, limiting our number of contemporaneous positive NP and saliva specimens. However, we overcame this by using saliva spiked with positive NP specimens and comparing the analytical performance using CT values as a more precise measure.

Other authors have found comparable results using the CDC’s version of the SARS-CoV-2 using unprocessed saliva in symptomatic adult individuals.^10^ Chong et al. found testing of saliva collected from children to be less sensitive than testing NP specimens in viral transport medium. We initially obtained similar results when testing unprocessed saliva, however, when we collected saliva from children with the SpectrumDNA saliva collection kit, we found comparable sensitivity as measured by CT values from NP and saliva specimens.

In conclusion, our study findings indicate that saliva collected from COVID-19 infected symptomatic and asymptomatic children that is mixed with SpectrumDNA saliva stabilizing solution yields clinically comparable PCR results compared to NP specimens. Use of saliva SARS-CoV-2 testing will enable greater access to sensitive PCR testing and may be advantageous for “back to school” testing programs for children.

## Data Availability

The data supporting the study findings are available within the paper.

## ACKNOWLEDGEMENTS

We are grateful to the Emergency Department Research Team who consented and enrolled all of the patients into this study; Michael Taylor, Christina Dollar Adebola Owolabi, Kara Hom, Ben Parrish and Sanyukta Deshmuk.

## AUTHORSHIP CONTRIBUTIONS

MD, JC designed the research, analyzed and interpreted data and wrote the manuscript. RD, JS designed the research and edited the manuscript. BT, CR, MM, JG, ME, MMc carried out the described studies. All authors reviewed and approved the final version of the manuscript.

